# Analytical Validation and Clinical Implementation of MedGenome HRD Assay:A Large Comparative Study Against Commercial HRD Assays in Ovarian Cancer

**DOI:** 10.64898/2026.11.02.26367052

**Authors:** Syed Muqlisur Rehman, Rohan Shetty, Coral Miriam Magdalene, Madhusmita Panda, Jammula Sri Ganesh, Ravi Gupta, Sakthivel Murugan, Vedam L. Ramprasad, Boman N Dhabar, Shekar Patil, Akhil Kapoor, Shyam Aggarwal, Tushar Patil, Ananth Pai, Krishna Prasad, Sourav Kumar Mishra, Vishwanath Sathyanarayanan, Raja Pramanik, Anuj Gupta, Bipinesh Sansar

**Affiliations:** Department of Molecular Oncopathology, MedGenome Labs Limited., Bangalore, Karnataka, India; Department of Surgical Oncology, Zulekha Institute of Oncology, Yenepoya (Deemed to be University), Mangalore, Karnataka, India; Department of Bioinformatics, MedGenome Labs Limited., Bangalore, Karnataka, India; Department of Medical Oncology, Fortis Hospital, Mulund, Mumbai, Maharashtra, India; Department of Medical Oncology, HCG Hospital, Bangalore, Karnataka, India; Department of Medical Oncology, Mahamana Pandit Madanmohan Malaviya Cancer Centre and Homi Bhabha Cancer Hospital, Tata Memorial Centre, Homi Bhabha National Institute, Varanasi, Uttar Pradesh, India; Department of Medical Oncology, Sir Ganga Ram Hospital, Delhi, India; Department of Medical Oncology, Sahyadri Speciality Hospital, Pune, Maharashtra, India; Department of Medical Oncology, Kasturba Medical College, Manipal, Karnataka, India; Department of Medical Oncology, Mangalore Institute of Oncology, Mangalore, Karnataka, India; Department of Medical Oncology, All India Institute of Medical Sciences, Bhubaneswar, Odisha, India; Department of Medical Oncologyfd, Apollo Hospital, Bannerghatta Road, Bangalore, Karnataka, India; Department of Medical Oncology, All India Institute of Medical Sciences, Delhi, India

**Author notes:** Corresponding authors: Syed Muqlisur Rehman, Madhusmita Panda.

**Keywords:** ovarian cancer, homologous recombination deficiency, genomic instability, *BRCA1/2*, mutation, high grade serous carcinoma

## Abstract

**Background:** Homologous recombination deficiency (HRD) is an important predictive biomarker for poly (ADP-ribose) polymerase inhibitor (PARPi) therapy in ovarian cancer (OC). However, real-world data on HRD prevalence and assay performance in Indian patients are limited. This study evaluated commercial HRD assays, then developed an in-house HRD assay and characterized HRD and *BRCA* alterations in a large real-world Indian OC cohort.

**Methods:** This retrospective study included 4,163 formalin-fixed paraffin-embedded (FFPE) OC samples tested between 2021 and 2025. HRD and *BRCA* status were initially assessed using the AmoyDx HRD Focus Panel (n=1,097) and SOPHiA DDM HRD Solution (n=470). Subsequently, 170 representative samples were used for the development and validation of an in-house HRD assay (MG_HRD), with external concordance assessed using the Myriad myChoice CDx assay in a subset of samples. Following validation, MG_HRD was implemented in routine clinical practice and applied to an independent cohort of 2,596 OC samples. HRD prevalence, *BRCA* mutation status, and clinicopathological associations were evaluated in all cohorts.

**Results:** Among 4,163 ovarian cancer samples, HRD positivity was observed in 52.4% of cases, and pathogenic/likely pathogenic *BRCA1/2* variants were identified in 23.6% of cases. Notably, 28.8% of tumors were *BRCA* wild-type but HRD-positive. *BRCA*-mutated tumors demonstrated significantly higher HRD genomic instability scores than *BRCA* wild-type tumors (p<0.001). HRD positivity was significantly enriched in high grade serous ovarian carcinomas. Remarkably, the *BRCA1* c.68_69del (p.Glu23ValfsTer17) founder mutation was detected in 14.2% (97/685) of cases among the pathogenic variants in our study. The MG_HRD assay showed excellent concordance with commercial assays, achieving 97.1% overall agreement, 98.3% negative percent agreement (NPA), and 96.4% positive percent agreement (PPA), with complete concordance observed in the externally evaluated Myriad subset.

**Conclusions:** This is the first Indian study to develop, validate, and implement an in-house HRD assay and evaluate HRD across three testing platforms in a large real-world OC cohort. The high prevalence of *BRCA* wild-type/HRD-positive tumors highlight the importance of comprehensive HRD testing, whereas the strong performance of MG_HRD supports its use as a reliable and cost-effective alternative for routine clinical practice.

## 1. Introduction

Homologous recombination deficiency (HRD) is a significant genomic instability phenotype resulting from impaired repair of DNA double-strand breaks through the homologous recombination repair (HRR) pathway^1^. This deficiency results in the accumulation of genomic scars, including loss of heterozygosity (LOH), telomeric allelic imbalance (TAI), and large-scale state transitions (LST), which together contribute to tumorigenesis^2^. HRD is highly prevalent in ovarian cancer (OC), particularly in high-grade serous carcinoma (HGSC), which is a common histological subtype, comprising 70% of OC cases^2,3^. HRD has become an important predictive biomarker for response to platinum-based chemotherapy and poly (ADP-ribose) polymerase inhibitors (PARPi) in OC. PARPi functions through the principle of synthetic lethality, where the inhibition of single-strand DNA repair results in the accumulation of DNA damage, which is lethal in HR-deficient tumors^4^.

Approximately 50% of HGSC cases exhibit HRD, either due to pathogenic *BRCA1/2* mutations [*BRCA1/2*_mut], which constitutes approximately 20-25% or through genomic scar markers such as LOH, TAI, and LST^3^. The HRD genomic scar score is calculated based on the quantification of these genomic scars^1^. While *BRCA1/2* mutation testing remains a cornerstone of molecular diagnostics, it is increasingly recognized that a significant proportion of HRD-positive tumors are *BRCA1/2* wild-type (*BRCA1/2*_wt). This highlights the need for comprehensive testing strategies that integrate both *BRCA1/2* mutation analysis and genomic scar score-based HRD assessment^5^. Next-generation sequencing (NGS) technology plays a crucial role in identifying patients for targeted therapeutics by enabling comprehensive analysis of the genomic scars and molecular alterations that drive ovarian cancer^6^.

Several commercial and laboratory-developed NGS-based assays are currently available for HRD testing. Among these, the Myriad MyChoice CDx assay (Myriad Genetics, Salt Lake City, UT, USA) is a Food and Drug Administration (FDA)-approved companion diagnostic HRD assay for PARP inhibitor therapy. This assay integrates *BRCA1/2* mutation status with a genomic instability score (GIS) derived from the combined score of LOH, TAI, and LST, with a predefined cutoff of ≥42 to define HRD positivity^7,8^. Despite its clear predictive value, the widespread adoption of this assay faces challenges because of its high cost, long turnaround time, and limited accessibility due to centralized testing^6^.

To overcome the limitations of the Myriad MyChoice CDx assay, several alternative commercial HRD diagnostic assays have been developed, including the AmoyDx HRD Focus Panel, SOPHiA DDM HRD Solution, FoundationOne CDx, Leuven HRD, and SeqOne HRD assays. Although these platforms are designed with the common objective of determining HRD status, they differ considerably in assay design, including gene content, single nucleotide polymorphism (SNP) backbone, sequencing methodologies and bioinformatic algorithms^7,9,10^. The lack of harmonized methodologies highlights the need for scalable, reproducible, and locally deployable diagnostic solutions for HRD assessment in ovarian cancer^3,6^. Each of these HRD assays uses different genomic instability metrics and positivity thresholds for HRD classification^7^. For example, the AmoyDx HRD Focus Panel uses a Genomic Scar Score (GSS) threshold of ≥50; the SOPHiA DDM HRD Solution applies a Genomic Integrity Index (GII) threshold of ≥0; FoundationOne CDx primarily relies on genomic loss of heterozygosity (gLOH) with a cutoff of ≥16; Leuven HRD employs a GIS derived from combined genomic scar markers with a cutoff of >56; and SeqOne HRD uses a proprietary genomic instability threshold^7^. Among these commercial decentralized NGS-based assays, AmoyDx and SOPHiA provide the most practical and reliable alternatives by enabling in-house HRD assessment using standardized analytical workflows. Their high concordance with established reference assays supports their implementation in routine clinical HRD testing^10–13^. Therefore, in this study, we chose to perform these two commercial assays in our MedGenome laboratory along with our in-house developed HRD assay, designated as the MedGenome HRD (MG_HRD) assay, to evaluate the HRD status in OC samples.

In this retrospective study, we validated the analytical performance and feasibility of the MG_HRD assay by comparing it with two commercially available HRD assays: AmoyDx and SOPHiA. A limited subset of samples underwent external benchmarking using the Myriad myChoice CDx assay. As mentioned above, the HRD metrics of all these assays are different. Hence, to ensure consistency across platforms, these metrics were collectively referred to as the HRD genomic score (HRD_GS) in this study. The primary objectives of this study were to assess cross-platform agreement using concordance metrics and to evaluate the analytical performance of the MG_HRD assay. The secondary objectives were to determine the prevalence of HRD, alterations in *BRCA1/2*, and the status of HRD_GS across the three assays. Furthermore, we investigated the association between HRD status with tumor content, and histological subtypes across a large, independent ovarian cancer cohort for all three assays. Through this comprehensive evaluation, we aimed to investigate the applicability and scalability of MG_HRD testing for the molecular diagnosis of ovarian cancer.

## 2. Methodology

### 2.1. Study Design

In this study, a large independent clinical cohort of 4163 patients with OC was analyzed for HRD testing using three distinct assays. These included two commercially available assays (AmoyDx, SOPHiA) and one internally developed assay (MG_HRD). Among these, 1097 patients (26.4%) were assessed using the AmoyDx (assay 1), 470 patients (11.3%) using the SOPHiA (assay 2), and 2596 patients (62.4%) using the MG_HRD (assay 3). A small subset of samples was analyzed using the SOPHiA assay (n = 14), which yielded inconclusive HRD results. Therefore, these samples were excluded from downstream analysis, resulting in 470 evaluable samples for the SOPHiA assay [**Figure 1A**].

**Figure 1:**
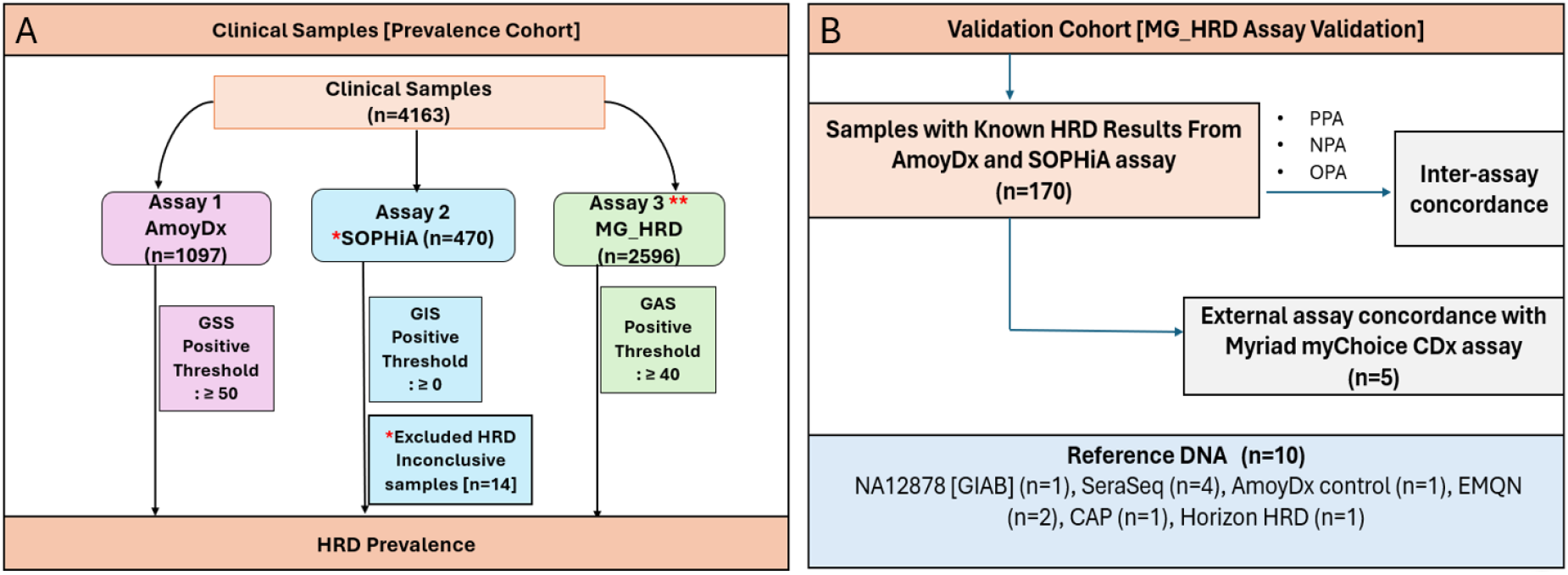
Workflow of the HRD analysis. A: Prevalence cohort of total samples; B: Validation cohort samples only. ** Following the successful analytical validation of the MG_HRD assay, clinical samples were analysed using this assay.

The analytical performance of the MG_HRD assay was assessed using an internal validation cohort of 170 OC samples. These samples were selected from the prevalence cohort of 1567 samples with known HRD results from the AmoyDx [157**/**1097] and SOPHiA [13/470] assays. Additionally, commercially available reference DNA samples (n = 10) with known HRD_GS and *BRCA1/2* mutation statuses were included in the validation process to optimize the analytical workflow of the assay. The reference standards comprised NA12878 [GIAB] (n=1), SeraSeq (n=4), AmoyDx control (n=1), EMQN (n=2), CAP (n=1), and Horizon HRD (n=1). The MG_HRD validation cohort (n=170) was stratified into four groups based on the status of HRD_GS and *BRCA1/2* mutations: *BRCA1/2*_mut/HRD_GS+(n=46), *BRCA1/2*_mut/HRD_GS-(n=14), *BRCA1/2*_wt/HRD_GS+(n=52), and *BRCA1/2*_wt/HRD_GS- (n=58). To further evaluate the concordance of the assays with an external reference assay, five samples from the validation cohort were evaluated using the FDA-approved Myriad myChoice CDx Assay. To represent the molecular subgroups as defined above, four samples were chosen at random, ensuring that one sample was taken from each subgroup. Additionally, one sample that failed the AmoyDx library quality control (QC) criteria was included because of its high tumor cellularity. This QC-failed sample was analyzed using the MG_HRD assay to assess analytical performance and was exclusively included in the external validation. However, this sample was not considered a part of the assay validation or overall prevalence cohort [**Figure 1B**].

Formalin-fixed paraffin-embedded (FFPE) ovarian tissue samples were analyzed at MedGenome Labs, Bangalore, India, between January 2021 and May 2025. This single-center retrospective study was approved by the Institutional Ethics Committee (IEC No. EC/NEW/INST/2022/3194; Proposal No. 02/2023). The study was conducted in accordance with the ethical principles of the Declaration of Helsinki and the ICMR’s National Ethical Guidelines for Biomedical and Health Research Involving Human Participants. All patient information was de-identified prior to analysis to ensure confidentiality. The inclusion criteria encompassed FFPE tumor tissue with adequate tumor content, and a clinical request for HRD testing. The clinicopathological variables evaluated in this study included patient age, disease stage, histological subtypes, and presence of metastasis.

### 2.2. Processing of FFPE Tumor Tissue Samples and DNA extraction

FFPE ovarian tumor tissue samples underwent initial histopathological assessment following hematoxylin and eosin (H&E) staining. Samples with a tumor content (TC) of ≥30% were preferentially selected for HRD testing based on the minimum TC requirement of the AmoyDx and SOPHiA assays. Cases with TC <30% were also included, provided that TC was ≥20%, consistent with the minimum requirement of the in-house MG_HRD assay. In cases where the TC was insufficient, an additional FFPE block from the same patient was reviewed. Subsequently, tissue sections of 10 µm thickness were sliced using a microtome, and the volume of the FFPE curls was adjusted based on the TC present in each block. The selected sections were then deparaffinized prior to molecular processing.

Genomic DNA was extracted from FFPE tissue sections using the QIAamp DNA FFPE Tissue Kit (Qiagen, Cat No. 56404, Germantown, MD, USA) as per the manufacturer’s protocol. The concentration and purity of the isolated DNA were assessed via fluorometric quantification using Qubit (Thermo Fisher Scientific, Waltham, MA, USA), while the fragment size distribution was evaluated using the TapeStation system (Agilent Technologies, Santa Clara, CA, USA).

### 2.3. HRD testing

#### 2.3.a. AmoyDx HRD Focus Panel

HRD profiling of assay1 samples [n=1097] was performed using the AmoyDx HRD Focus Panel (AmoyDx, Xiamen, China), according to the manufacturer’s guidelines [Version: B1.0]^14^. Genomic DNA extracted from FFPE tumor tissue (100–200 ng) was used for library preparation based on Halo-shape Annealing and Defer-Ligation Enrichment (HANDLE) technology, followed by target enrichment, library amplification, purification, and quantification. Libraries were sequenced on a NovaSeq 6000 platform (Illumina, San Diego, CA). Sequencing reads were aligned with the human reference genome (hg19). Variants were called to identify P/LP single nucleotide variants (SNVs), small insertions and deletions (InDels), and splice site mutations in the *BRCA1/2* genes.

In addition to mutation detection, the assay quantifies genomic instability using approximately 24,000 genome-wide SNP markers, enabling the inference of allele-specific copy number alterations through normalized read depth and B-allele frequency data^6,8^. Bioinformatic data analysis was conducted through the AmoyDx NGS Data Analysis System (ANDAS) (Amoy Diagnostics Co., Ltd.)^14^. The Genomic Scar Score (GSS) was calculated using a machine-learning (ML) algorithm that integrates chromosomal copy-number alterations (CNAs) based on their length, type, and location^6^. The predefined HRD GSS threshold for positivity was set at GSS ≥50. Samples with a GSS ≥50, and/or those harboring pathogenic *BRCA1/2* variants were classified as HRD-positive.

#### 2.3.b. SOPHiA DDM^TM^ HRD Solution

HRD profiling of assay2 samples [n=470] was performed using the SOPHiA HRD™ solution (SOPHiA GENETICS, Lausanne, Switzerland) according to the manufacturer’s instructions^15^. This assay integrates targeted NGS of multi-HRR genes with low-pass whole genome sequencing (LP-WGS) for a genome-wide assessment of genomic instability. Genomic DNA extracted from FFPE tumor tissue (50–100 ng) was used for library preparation using the SOPHiA GENETICS™ DNA Library Prep Kit III. Library quality and quantity were assessed using Qubit fluorometry (Thermo Fisher Scientific) and an Agilent Fragment Analyzer™ [Agilent Technologies], followed by sequencing on an Illumina NovaSeq 6000 platform.

The targeted HRR panel enabled the detection of SNVs, InDels, and CNVs, including alterations in *BRCA1/2*, whereas LP-WGS was performed at low coverage to capture genome-wide copy number profiles for genomic instability assessment^6^. Sequencing data were processed using the SOPHiA DDM™ platform, aligned to the human reference genome (hg19). Genome-wide coverage profiles were normalized and analyzed using a deep neural network-based algorithm to derive the Genomic Integrity Index (GII). The GII quantifies chromosomal aberration patterns, and the predefined positive threshold was GII ≥0. Samples exceeding the GII threshold and tumors with pathogenic *BRCA1* or *BRCA2* mutations were classified as HRD-positive. Samples not meeting predefined QC criteria for DNA input, sequencing depth, *BRCA* coverage, or inconclusive HRD score were excluded from the analysis.

#### 2.3.c. MG_HRD Assay

##### i. Design of MG_HRD Panel

To enable HRD assessment using our indigenous assay, we designed a custom panel with dual functionality: (a) genome-wide detection of copy number alterations using strategically distributed SNP markers, (b) identification of pathogenic variants in *BRCA1/2* and other homologous recombination repair (HRR) genes. The panel incorporates genome-wide SNP biallelic markers selected to ensure broad representation across diverse ethnic populations, together with a targeted gene panel comprising key HRR-associated genes and *CCNE1*. The targeted panel includes 17 genes: *ATM, BARD1, BRCA1, BRCA2, BRIP1, CDK12, CHEK1, CHEK2, FANCL, FGFR1, PALB2, PPP2R2A, RAD51B, RAD51C, RAD51D, RAD54L*, and *CCNE1*.

The genome-wide SNP backbone was selected from well-curated population reference databases, including the 1000 Genomes Project, GenomeAsia, gnomAD, and our internally curated SAS-ATLAS database, which comprises data from over 100000 samples. SNP selection was performed by adapting the methodology described by Timms et al^16^. Briefly, candidate SNPs were filtered to retain markers with a minor allele frequency (MAF) greater than 5%, low linkage disequilibrium and exclusion from regions associated with segmental duplications, problematic genomic regions, repetitive elements and low-mappability sequences. Using this strategy, approximately 46,000 genome-wide SNP markers were prioritized for panel design. The shortlisted SNPs were subsequently evaluated for probe design, and the optimal probes were selected based on simulated hybridization performance. The probes for both SNPs and HRR targets were densely and differentially tiled (3X-10X) for better capturing in the poor quality FFPE.

The coding regions (CDS) of *BRCA1/2* and other HRR genes were selected based on latest MANE (Matched Annotation from NCBI and EBI) and GENCODE gene models. Our design also covered 20 bases upstream and downstream of the exon-intron boundaries for the identification of splice site disease causing variants.

##### ii. Sample preparation and sequencing

Approximately 50–100 ng of DNA was used for targeted library preparation using a custom hybrid capture kit (Twist Library Prep Kit EF with AMP Mix cat#101058 Twist Bioscience, USA). Library quality was checked using a Qubit (Thermo Fisher Scientific, USA) and TapeStation (Agilent, USA). Libraries that met the quality standards were quantified, loaded at a concentration of 120 pM, and sequenced as 2 × 150 bp paired-end reads on an Illumina Novaseq X plus. The sequencing process adhered to the manufacturer’s protocol, employing the NovaSeq X Series 25B Reagent Kit 300Cyc (Cat No. 20104706), and achieved an average sequencing depth of approximately 250x for the analysis of somatic mutations.

##### iii. Analysis of sequencing data

The sequencing data undergoes an optimized workflow to determine the HRD status. Initially, this workflow assesses the quality of the reads and trims them as needed. After trimming, the reads were aligned to the hg38 reference genome and collapsed based on Unique Molecular Identifier (UMI) information to generate a consensus BAM file. Variant calling was then performed on this consensus BAM file for all targeted regions of HRR genes. Deleterious mutations were identified using a well-established mutation prioritization process and annotated with the comprehensive mutation annotation tool, VariMAT. This tool annotates mutations across various population and disease databases.

This assay evaluates the HRD status using a combined approach based on the genome aberration (GA) score, which is a quantitative measure of genomic alterations derived from high-quality ∼46K SNP markers and clinically significant variants from the *BRCA1/2* gene. This assay identifies high-quality SNVs, InDels, and splice site mutations and estimates allele-specific copy numbers based on their bi-allele frequency and coverage. The GA score (GAS) was calculated by considering different levels of copy number changes with a positivity threshold of ≥40. A sample is classified as HRD-positive if P/LP variants were detected in the *BRCA1/2* genes and/or the GAS ≥40. Additionally, this assay determines the copy number status of *CCNE1*, which was considered amplified if its copy number was ≥7.

### 2.4. Variant Classification

In the evaluation of HRD in ovarian cancer, current evidence suggests that only pathogenic or likely pathogenic (P/LP) variants in the *BRCA1/2* genes hold clinical significance. Consequently, although the SOPHiA and MG_HRD assays include multiple genes in the homologous recombination repair (HRR) panel, this study focused exclusively on *BRCA1/2* variants for HRD interpretation. The identified somatic *BRCA1/2* variants were classified according to the ASCO-CAP and AMP guidelines, categorizing them as pathogenic, likely pathogenic, or variants of uncertain significance (VUS)^17^. Nonsense, frameshift insertions/deletions, in-frame and canonical splice site variants classified as P/LP were grouped as pathogenic. These pathogenic variants were further verified using the ClinVar, Human Gene Mutation Database (HGMD) and BRCA Exchange databases, and only clinically significant variants were considered for downstream HRD status analyses. Variants of uncertain significance (VUS) in *BRCA1/2* genes were classified as negative for *BRCA* status within this HRD assessment framework.

In this study, we did not conduct germline testing; therefore, the *BRCA1/2* mutations identified could be of either germline or somatic origin. Mutations in *BRCA1/2* with a variant allele frequency (VAF) exceeding 40% were considered to infer the probability of germline origin.

### 2.5. Statistical Data Analysis and Visualization

Statistical data analysis was done by using Python (version 3.13.3)^18^. Python libraries, such as NumPy^19^, Pandas^20^, and SciPy^21^, were used for the statistical analysis of both descriptive and categorical analysis^22^. Descriptive statistics were used to summarize the distributions of sequencing QC parameters and HRD scores using the means, medians, and interquartile ranges (IQR). The Mann–Whitney U test was used to compare HRD scores between the *BRCA1/2*_mut and *BRCA1/2_*wt groups. Associations between HRD status and categorical clinicopathological variables, including tumor content and histological classification, were assessed using the chi-square test. Results were considered statistically significant if p < 0.05. Matplotlib^23^ and Seaborn^24^ Python libraries were used for the data visualization. All pathogenic *BRCA1/2* variants were illustrated using a lollipop plot generated via cBioPortal with the genome build hg38.

## 3. Results

### 3.1. Clinicopathological characteristics of patients

A total of 4163 FFPE OC samples from both prevalence and validation cohorts were retrospectively analyzed using three HRD testing platforms: AmoyDx (n = 1097), SOPHiA (n = 470), and MG_HRD (n = 2596). The clinicopathological characteristics of the total cases are summarized in **Table 1**. The median age at diagnosis was 55 years, and the mean age was 54.6 years. The cohort predominantly comprised patients with FIGO stage III disease (53.5%), followed by stage II (36.2%). High-grade serous carcinoma (HGSC) was the most frequently represented histological subtype (82.5%). Most FFPE specimens were obtained from primary tumor sites (76.4%), while 23.6% were derived from metastatic sites, primarily pelvic lymph nodes and peritoneal nodules, followed by liver.

**Table 1:** Clinicopathological Characteristics of Patients.

| **Characteristics** | **Assay Types** |  |  | **Total**<br>**N (%)** |
|---|---|---|---|---|
|  | **AmoyDx**<br>**N (%)** | **SOPHiA**<br>**N (%)** | **MG_HRD**<br>**N (%)** |  |
| Patients | 1097 (26.4%) | 470 (11.3%) | 2596 (62.4%) | 4163 (100%) |
| **Age at diagnosis** |  |  |  |  |
| Median | 55 | 55 | 55 | 55 |
| Mean ± SD | 54.3 ± 11.2 | 54.7 ± 12 | 54.7 ± 10.9 | 54.6 ± 11.1 |
| **Stage** |  |  |  |  |
| Stage: I | 73 (6.7%) | 39 (8.3%) | 28 (1.1%) | 140 (3.4%) |
| Stage: II | 344 (31.4%) | 195 (41.5%) | 967 (37.2%) | 1506 (36.2%) |
| Stage: III | 604 (55.1%) | 204 (43.4%) | 1421 (54.7%) | 2229 (53.5%) |
| Stage: IV | 76 (6.9%) | 32 (6.8%) | 180 (6.9%) | 288 (6.9%) |
| **Histological Classification** |  |  |  |  |
| High grade serous carcinoma | 995 (90.7%) | 411 (87.4%) | 2028 (78.1%) | 3434 (82.5%) |
| Low grade serous carcinoma | 28 (2.6%) | 14 (3.0%) | 274 (10.6%) | 316 (7.6%) |
| Endometrioid carcinoma | 18 (1.6%) | 16 (3.4%) | 97 (3.7%) | 131 (3.1%) |
| Clear cell carcinoma | 19 (1.7%) | 11 (2.3%) | 87 (3.4%) | 117 (2.8%) |
| Mucinous carcinoma | 11 (1.0%) | 2 (0.4%) | 33 (1.3%) | 46 (1.1%) |
| Carcinosarcoma | 4 (0.4%) | 1 (0.2%) | 12 (0.5%) | 17 (0.4%) |
| Serous_unknown grade | 22 (2.0%) | 15 (3.2%) | 65 (2.5%) | 102 (2.5%) |
| **Primary v/s Metastatic** |  |  |  |  |
| Primary | 896 (81.7%) | 394 (83.8%) | 1892 (72.9%) | 3182 (76.4%) |
| Metastatic | 201 (8.3%) | 76 (16.2%) | 704 (27.1%) | 981(23.6%) |

### 3.2. Development and Analytical Validation of the MG_HRD Assay

To evaluate analytical performance, the MG_HRD assay was validated using reference standards and clinical samples with known HRD status of AmoyDx [n=157] and SOPHiA [n=13] assays. Higher concordance was observed between AmoyDx/SOPHiA and MG_HRD assays for these 170 OC samples, yielding 96.39% Positive Percentage Agreement (PPA), 98.3% Negative Percent Agreement (NPA), and 97.05% Overall Percent Agreement (OPA). Of the 170 samples, 165 had the same HRD status, whereas only five were discordant. Among these five discordant cases, all were concordant with *BRCA1/2*_wt results, while only discordant at HRD_GS values. Four out of five samples were HRD_GS-positive by AmoyDx (GSS ≥50) but HRD_GS-negative by MG_HRD (GAS <40), whereas one sample showed the reverse result. Of these four discordant cases, two had borderline scores near the respective assay thresholds (GSS: 50.1, GAS: 32; GSS:66.1, GAS: 39). This may reflect differences in assay-specific scoring algorithms, score calculation methods, and positivity cutoffs. The contingency table summarizing the comparison between the MG_HRD assay and the alternate assays is presented below, along with the calculated agreement metrics [**Table 2A**].

**Table 2A:** Comparison of HRD status determined by the MG_HRD assay with results obtained from the SOPHiA DDM HRD solution and the AmoyDx HRD Focus Panel.

| **MG_HRD Assay** | **AmoyDx / SOPHiA** |  | **PPA: 96.4%**<br>**NPA: 98.3%**<br>**OPA: 97.1%** |
|---|---|---|---|
|  | **Positive (n=111)** | **Negative (n=59)** |  |
| **Positive (n=108)** | 107 | 1 |  |
| **Negative (n=62)** | 4 | 58 |  |

In addition, four selected samples from the validation cohort [n=170] and sample_5 were analyzed externally using the Myriad myChoice HRD assay. The detailed results from AmoyDx, the MG_HRD assay, and Myriad myChoice are summarized in **Table 2B**. Notably, the HRD status classification for all five samples demonstrated 100% concordance between the MG_HRD and outsourced Myriad myChoice assays, further supporting the analytical reliability of the MG_HRD method. Although Myriad did not specify the HRD_GS for sample_2, the *BRCA* status for this sample was positive, resulting in overall HRD positivity. In contrast, the same sample was able to provide both HRD_GS and *BRCA* status by the MG_HRD assay, thus achieving concordance. The MG_HRD assay also showed 100% concordance with the AmoyDx assay in four samples. Interestingly, sample_5 failed at the library QC level for the AmoyDx assay but showed a positive HRD status [both HRD_GS and *BRCA* status] for our MG_HRD assay and the Myriad assay. The overall results showed that the MG_HRD assay was highly concordant with both the AmoyDx and Myriad assays.

**Table 2B:** Comparison of HRD status across the AmoyDx, MG_HRD assay, and Myriad results.

| Sample ID |  | S_1 | S_2 | S_3 | S_4 | S_5 |
|---|---|---|---|---|---|---|
| Tumor Content |  | 90% | 25% | 70% | 75% | 90% |
| AmoyDx Assay | *BRCA* status | Negative | Positive | Positive | Negative | Lib. QC Fail |
|  | HRD_GS status | 1.8 | 14.3 | 83.6 | 98.6 | Lib. QC Fail |
|  | HRD status | Negative | Positive | Positive | Positive | NA |
| MG_HRD Assay | *BRCA* status | Negative | Positive | Positive | Negative | Positive |
|  | HRD_GS status | 4 | 0 | 74 | 84 | 52 |
|  | HRD status | Negative | Positive | Positive | Positive | Positive |
| Myriad MyChoice CDx Assay | *BRCA* status | Negative | Positive | Positive | Negative | Positive |
|  | HRD_GS status | 3 | Unable to analyze | 46 | 84 | 52 |
|  | HRD status | Negative | Positive | Positive | Positive | Positive |

#### Limit of detection (LoD) and precision of the MG_HRD assay

Using FFPE samples, the assay was evaluated across a range of tumor content proportions (10–90%). Reliable detection of changes was achieved in samples containing as little as 20% tumor content. HRD scores remained consistent across varying DNA input amounts (200, 100, 50, and 30 ng), with reduced consistency observed only at 10 ng. Based on these findings, 30 ng was established as the optimal minimum DNA input requirement for the assay. The limit of detection (LoD) demonstrated 100% concordance at a 1% alternate allele frequency. The MG_HRD assay was evaluated using the same set of samples across multiple Illumina sequencing platforms, with different flow cell types, and by different operators on separate days. The MG_HRD assay results demonstrated 100% concordance across all testing conditions, indicating excellent reproducibility and robustness.

### 3.3. Sequencing performance Across the Platforms

Analysis of QC metrics revealed overall a good sequencing performance for all the three different assays. In the AmoyDx cohort, median alignment rate was 98.9% (IQR: 98.6-99.2%) and median effective depth of *BRCA* and SNP was 2498× and 472× respectively. The SOPHiA cohort achieved the high target coverage with a median *BRCA* coverage and panel coverage percentile of 100% and 99.9%, respectively. The median percentage of total fragmentation and whole-genome fragments were 15.0% and 70.4%, respectively. In the MG_HRD cohort, median coverage values for *BRCA1* and *BRCA2* were 100.0% and 95.3%, respectively, with a median backbone coverage of 90.5%. This assay obtained a median panel depth of 251x and 80% on target reads, allowing for adequate coverage to accurately assess both *BRCA1/2* genes and genome-wide backbone regions. Despite differences in platform-specific QC metrics and sequencing designs, all assays consistently indicated greater coverage and robust sequencing quality for both *BRCA* and HRD analysis.

### 3.4. Distribution of HRD parameters

The HRD parameters included *BRCA1/2* mutations and HRD_GS. Across all three assays, approximately half of the tumors were classified as HRD-positive (*BRCA1/2* mutation and/or a positive HRD_GS). The HRD-positivity ranges from 44.7% (210/470) with SOPHiA to 54.8% (601/1097) with AmoyDx and 52.8% (1370/2596) with MG_HRD [**Figure 2A**]. Overall, 52.4% (2181/4163) of tumors were HRD-positive. Among the HRD-positive subgroups, *BRCA1/2*_wt/HRD_GS-positive tumors constituted the largest subgroup overall (28.8%), with frequencies of 31.6%, 20.9%, and 29.0% for AmoyDx, SOPHiA, and MG_HRD assay, respectively. *BRCA1/2*_mut/HRD_GS-positive and *BRCA1/2*_mut/HRD_GS-negative tumors accounted for 16.7% and 6.9%, respectively of the overall cohort, while *BRCA1/2*_wt/HRD_GS-negative [HRD-negative] subgroup comprised 47.6% [**Table S1**]. Notably, the distribution of the molecular subgroups was broadly consistent between AmoyDx and MG_HRD, whereas SOPHiA showed a relatively lower proportion of *BRCA1/2*_wt/HRD_GS-positive tumors and a higher proportion of HRD-negative tumors [**Figure 2B**]. Pathogenic *BRCA1/2* mutations were detected at comparable frequencies, ranging from 23.2% to 23.8% with an overall frequency of 23.6% [983/4163]. Whereas the HRD_GS-positivity was observed at 45.4% [1892/4163] of the total samples by using assay-specific thresholds [**Table S2**].

**Figure 2:**
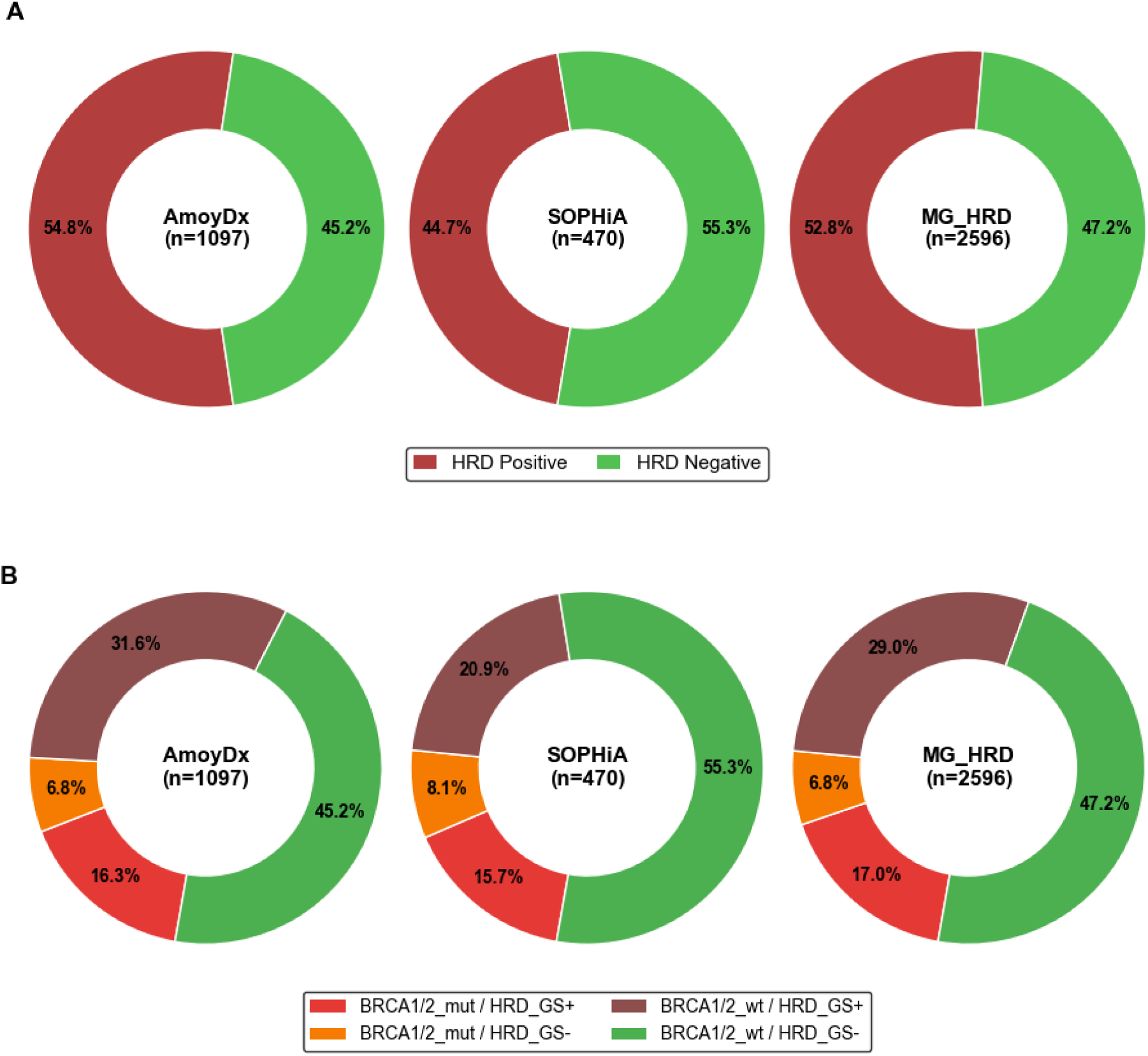
Distribution of HRD and *BRCA* status; A: Distribution of Overall HRD status; B: Combined distribution of *BRCA* and HRD_GS Status across three assays.

### 3.5. Association between *BRCA1/2* Mutation and HRD_GS status

Across the three assay platforms, *BRCA1/2*_mut tumors demonstrated significantly elevated HRD genomic scores than *BRCA1/2*_wt tumors, with median (IQR) score of 90.3 (23.15 to 98.78) vs 22.2 (4.15 to 91.40) for AmoyDx, 3.0 (-0.92 to 7.20) vs -4.1 (-8.98 to 1.42) for SOPHiA, and 57.0 (31 to 70) vs 28.0 (5 to 57) for MG_HRD assay, respectively (all *P* < 0.0001; Mann-Whitney U test) [**Figure 3**]. Consistent with these findings, HRD_GS-positivity was significantly higher among *BRCA1/2* mutated than wild-type tumors, occurring in 70.5% vs 41.2% of AmoyDx, 66.1% vs 27.4% of SOPHiA, and 71.5% vs 38.0% of MG_HRD cases, respectively (all *P* < 0.0001; Chi square test) [**Table S3**].

**Figure 3:**
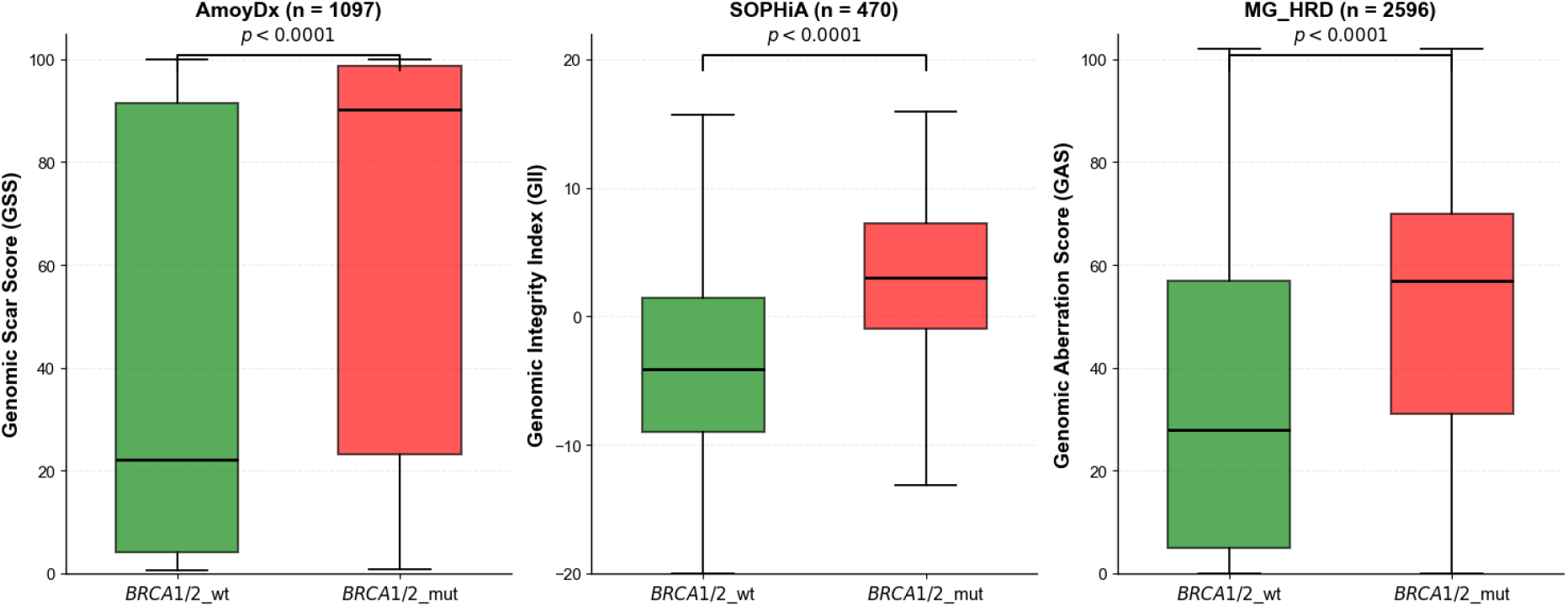
Association of *BRCA1/2* Mutation Status with HRD_GS Across Assay Types. p-values were calculated with a Mann–Whitney U test. The median score distributions were represented by horizontal lines in the plots. X-axis represents *BRCA* mutation status, while Y-axis represents HRD scores. Y-axis scale ranges from 0 to 100 for AmoyDx and MG_HRD assay; while -20 to +20 for SOPHiA assay. HRD matrices were different for each assay; GSS for AmoyDx; GII for SOPHiA; GAS for MG_HRD. Abbreviations: GSS: Genomic Scar score; GII: Genomic Instability Index; GAS: Genomic Aberration Score; *BRCA1/2*_mut: *BRCA1/2* mutant; *BRCA1/2*_wt: *BRCA1/2* wild-type

### 3.6. Association between TC and HRD Status across the assay types

Across all three HRD assays, higher tumor content was significantly associated with increased HRD positivity (p < 0.0001), with rates rising from 28.5% to 61.6% for AmoyDx, 21.2% to 60.0% for SOPHiA, and 33.2% to 61.8% for MG_HRD across the <30%, 30–60%, and >60% tumor content groups. Overall, HRD positivity increased with tumor content for all three assays, from lowest at TC <30% to highest at TC >60% [**Figure 4; Table S4**].

**Figure 4:**
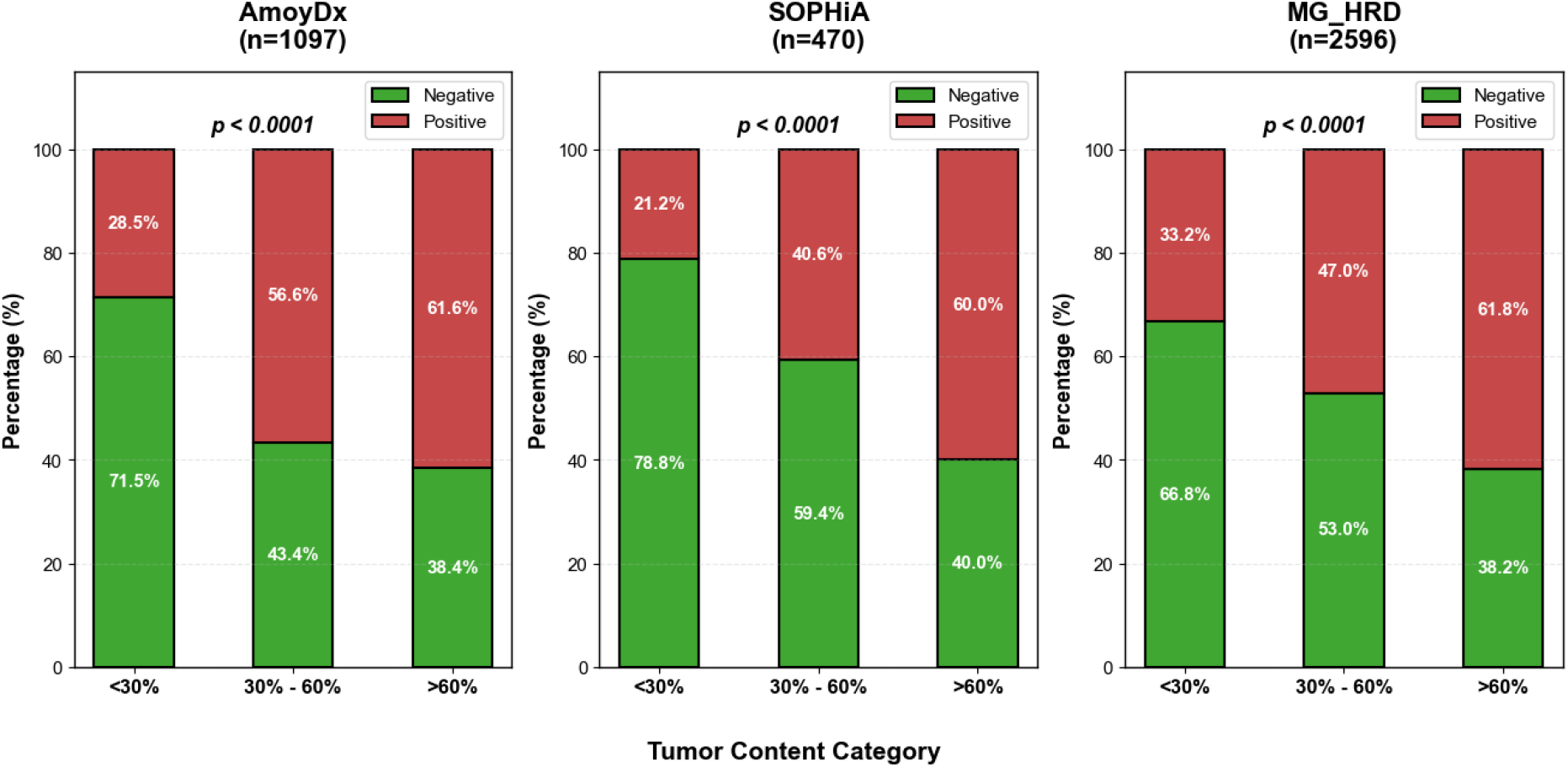
Association of Tumor Content with HRD Status Across Three Assay Platforms. p-values were calculated with chi-square test.

### 3.7. Association between histological subtype and HRD status

HGSC represented the predominant subtype (n = 3434/4163), exhibiting a significantly higher percentage of HRD-positive tumors (60.2%). In contrast, non-HGSC subtypes demonstrated substantially lower HRD positivity rates, such as low-grade serous (3.8%), mucinous (2.2%), clear cell (13.7%), and endometrioid carcinomas (26.7%). Carcinosarcoma had a relatively high HRD positivity rate of 52.9%. Serous tumors with unknown grades had an intermediate HRD positivity rate of 41.2%. The Chi-square test indicated a strong significant association between the histologic subtype and HRD status (p <0.0001). When HRD status was compared between the HGSC and non-HGSC groups, excluding serous tumors of unknown grade (n=102), a highly statistically significant association was observed between these groups (p <0.0001). The exclusion of serous unknown grade cases was performed to minimize ambiguity classification [**Table 3**].

**Table 3:** Association between Histological Subtype and HRD Status in Ovarian Cancer.

| Histologic subtype<br>(n = 4163) | Total (n) | HRD positive<br>n (%) | HRD negative<br>n (%) | p-value |
|---|---|---|---|---|
| HGSC | 3434 | 2066 (60.2%) | 1368 (39.8%) |  |
| Low-grade serous carcinoma | 316 | 12 (3.8%) | 304 (96.2%) | <0.0001 |
| Endometrioid carcinoma | 131 | 35 (26.7%) | 96 (73.3%) |  |
| Clear cell carcinoma | 117 | 16 (13.7%) | 101 (86.3%) |  |
| Mucinous carcinoma | 46 | 1 (2.2%) | 45 (97.8%) |  |
| Carcinosarcoma | 17 | 9 (52.9%) | 8 (47.1%) |  |
| Serous unknown grade | 102 | 42 (41.2%) | 60 (58.8%) |  |
| **HGSC vs non-HGSC (n = 4061)** |  |  |  |  |
| HGSC | 3434 | 2066 (60.2%) | 1368 (39.8%) |  |
| Non-HGSC | 627 | 73 (11.6%) | 554 (88.4%) | <0.0001 |

p-values were calculated with Chi-square test. HGSC: High grade serous carcinoma.

### 3.8. Mutation spectrum of *BRCA1* & *BRCA2 Genes*

Of the 4163 patients, *BRCA1/2* pathogenic mutations were observed in 983 patients [23.6%]. A total of 1002 pathogenic *BRCA1/2* variants were identified in 983 patients, comprising 693 *BRCA1* (16.6%) and 309 *BRCA2* (7.4%) variants. Among these, 18 patients harbored co-occurring pathogenic variants in both *BRCA1* and *BRCA2* [**Table S5**]. Recurrent pathogenic variants were classified as the variants that were detected with a frequency of ≥ 10 for *BRCA1* and ≥ 5 for *BRCA2*. Lollipop plots were generated to illustrate the distribution of pathogenic *BRCA1/2* variants [**Figure 5**].

**Figure 5:**
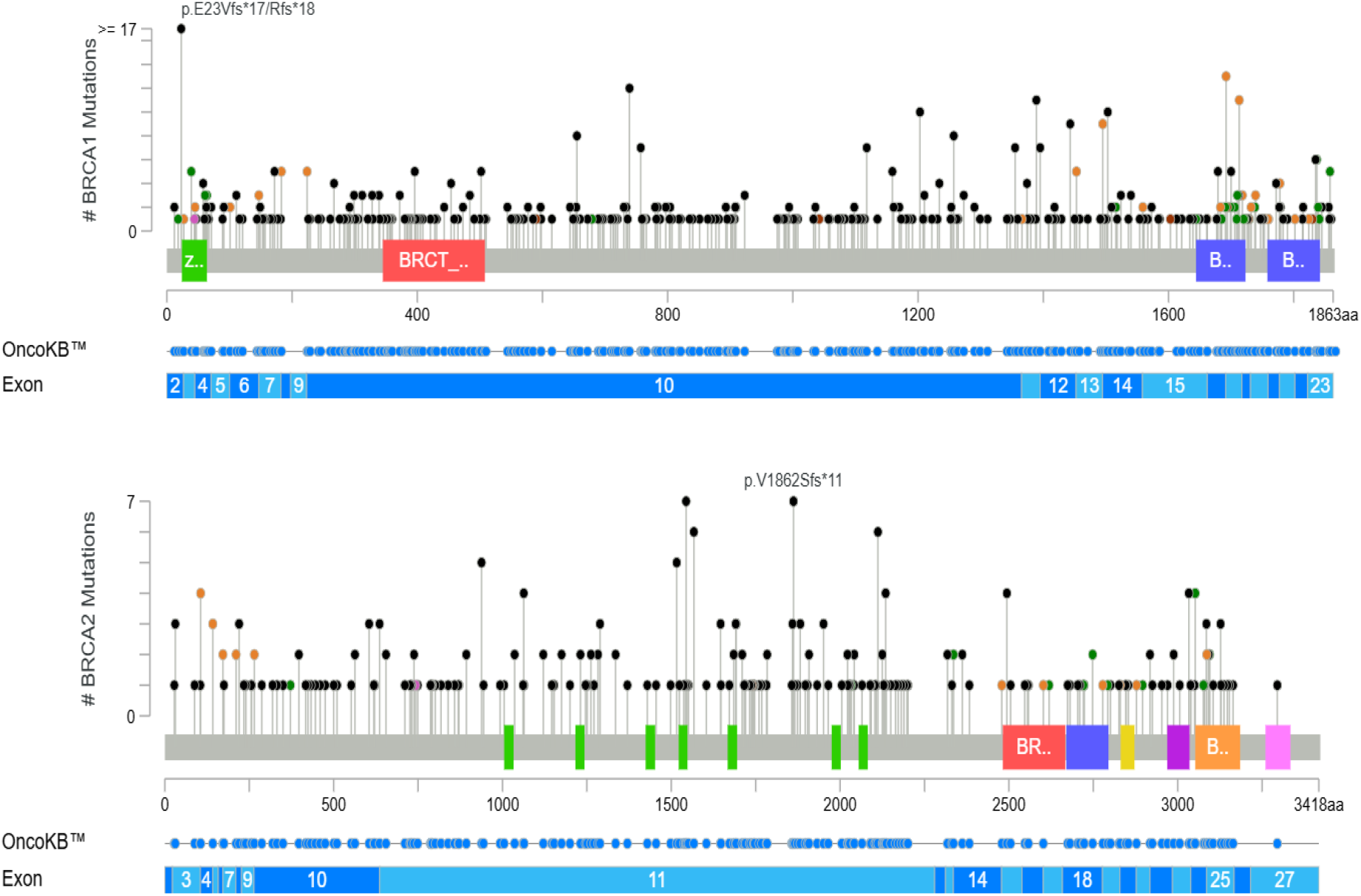
The lollipop plot illustrates the distribution of *BRCA1* and *BRCA2* pathogenic variants across the three HRD assays. The diagrams provide a linear representation of the BRCA1/2 protein domains along the x-axis. Each variant is depicted by an individual lollipop, with the height of the stick reflecting the mutation frequency on the y-axis. The circles are color-coded according to the variant type. Black circles indicate truncating variants, which include frameshift deletions/insertions and InDels. In-frame, splice-site, and missense variants are indicated by red, yellow, and green circles, respectively. Purple circles denote the other variants, including silent mutations. *BRCA1* domains: green, C3HC4-type RING finger; red, Serine rich domain associated with BRCT; blue, BRCT (BRCA1 C-terminal) domain. *BRCA2* domains: green, *BRCA2* repeat regions; red, *BRCA2* helical; blue, *BRCA2* oligonucleotide binding (OB) domain 1; orange, *BRCA2* OB3. This plot was generated using the Mutation Mapper tool available on cBioPortal. https://www.cbioportal.org/mutation_mapper.

In *BRCA1*, of the 693 pathogenic variants, the majority were truncating mutations (n = 561), followed by splice-site (n = 79), missense (n = 45), in-frame (n=6), and silent mutations (n = 2). *BRCA1* c.68_69del (p.Glu23ValfsTer17), the Ashkenazi founder mutation was the most recurrent variant, identified in 97 cases (14%), followed by the splice-site variant c.5074+1G>T (n=12). Additional recurrent variants included c.2214dup (p.Lys739Ter), c.4165_4166del (p.Ser1389Ter), and c.5138-2del. Similarly, *BRCA2* exhibited a high frequency of truncating variants (n = 267 out of 309), along with splice-site (n = 21), missense (n = 18), in-frame (n=1), and silent mutations (n = 2). The notable recurrent alterations observed in *BRCA2* included c.5583dup (p.Val1862SerfsTer11) and c.4631del (p.Asn1544ThrfsTer24), each detected in seven cases, followed by c.6332dup (p.Arg2112Efs*17) and c.4695_4698dup (p.Leu1567AspfsTer9), each detected in six cases. The distribution of *BRCA1/2* variants was enriched within functionally important domains. *BRCA1* variants are predominantly localized to the N-terminal RING finger and C-terminal BRCT domains, whereas *BRCA2* variants are distributed across the BRC repeat regions. Within the cohort, most pathogenic variants were identified in exon 10 of *BRCA1* and exon 11 of *BRCA2*.

### 3.9. *CCNE1* Amplification Status of MG_HRD Assay

Among the three HRD assays, *CCNE1* amplification was evaluated only using the MG_HRD assay. Of the 2596 patients, *CCNE1* amplification (i.e., ≥ seven copies) was detected in 181 (7%) cases. Among HRD-positive tumors, 87/1370 (6.4%) were *CCNE1*-amplified, versus 94/1226 (7.7%) of HRD-negative tumors. Overall, *CCNE1* amplification was significantly higher in *BRCA1/2*_wt cases, with a prevalence of 8.3% irrespective of GAS status, compared with *BRCA1/2*_mut cases (p < 0.0001) [**Table S6**].

## 4. Discussion

HRD testing in ovarian cancer has emerged as a crucial predictive biomarker, particularly for guiding therapeutic strategies involving PARP inhibitors. The FDA-approved CDx Myriad myChoice test is frequently employed for this purpose; however, for Indian patients it is costly, necessitates stringent quality standards, and involves prolonged result turnaround times, thereby limiting accessibility for patients^6,25^. Therefore, the development of more affordable and reliable in-house HRD assays are needed, which can be implemented in standard molecular laboratories that could enhance patient access, reduce costs, and expedite treatment decision-making. Various studies have described laboratory-developed HRD assays using different gene panels, HRD score thresholds, bioinformatic pipelines, and sequencing platforms^3,13^. Consistent with this, the recent AMP/ACCC/CAP joint consensus recommendations identified substantial heterogeneity in current commercial and laboratory-developed HRD assays regarding biomarker selection, scoring methods, validation strategies, and reporting practices^26^. These findings emphasize the need for rigorously validated and standardized in-house HRD assays to facilitate reliable clinical implementation and broader adoption in routine practice.

In this retrospective study, we present our experience of implementing an in-house developed HRD testing in our diagnostic laboratory. We assessed the feasibility, reliability and reproducibility of our MG_HRD assay by comparing results from the commercially available diagnostic assays [AmoyDx and SOPHiA] along with the reference assay, Myriad MyChoice CDx. We observed that the MG_HRD assay showed a complete concordance (100%) rate of HRD status with the AmoyDx assay among four samples and the outsourced Myriad myChoice CDx assay among five samples, which is similar to the reported studies^12,27^. High concordance was observed for HRD_GS status, *BRCA1/2* mutation status, and overall HRD classification, defined by the combined assessment of HRD_GS and *BRCA1/2* alterations. This result demonstrates that the MG_HRD assay can be used as a reliable and potentially more accessible alternative to the Myriad myChoice CDx for HRD assessment in ovarian cancer patients.

Previous studies have demonstrated a high level of agreement between different HRD assays and the Myriad assay. Fumagalli C et al. reported that AmoyDx HRD Focus Panel showed an overall percent agreement (OPA) of 87.8% (65/74 tumors) compared with MyChoice CDx^3^. Similarly, Wehn AK et al. reported an OPA of up to 96% (n=198), with a PPA of 95.2% and an NPA of 96.8% for Illumina’s TruSight Oncology 500 compared to MyChoice CDx^28^. The ENGOT European HRD initiative further developed the “Leuven” HRD assay as an academic alternative, demonstrating 91% overall agreement, with PPA and NPA of 94% and 86%, respectively, in 468 ovarian cancer samples from the PAOLA-1/ENGOT-ov25 trial^29^. The concordance of HRD status between the standardized Myriad assay (custom panel) and MyChoice® CDx test was 97.1% (499/514), according to Denkert et al^30^. In comparison, our MG_HRD assay showed the same concordance rate with an OPA of 97.1% (165/170), similar to the study conducted by Denkert et al^30^. A comparatively higher concordance of PPA (96.4 %) and NPA (98.3 %) was observed in the MG_HRD assay than in other studies^3,28,29^. These findings highlight the strong performance of the MG_HRD assay and its high degree of agreement with orthogonal platforms, including the AmoyDx HRD Focus and SOPHiA HRD assays, supporting its reliability for clinical implementation.

Our study indicated that the proportion of HRD-positive ovarian cancer cases ranged from 45% to 55% across the three evaluated assays. Specifically, the percentages were 44.7% for SOPHiA, 52.8% for MG_HRD and 54.8% for AmoyDx. These findings are consistent with previously published data. For instance, Guarischi-Sousa et al. reported HRD positivity rates of 45.9% and 65% using SOPHiA and AmoyDx assays, respectively^6^. Similarly, Fumagalli et al. observed 48.8% HRD-positive tumors using the AmoyDx assay, while data from The Cancer Genome Atlas (TCGA) indicated an approximate HRD prevalence of 50% among high-grade serous ovarian carcinoma (HGSC) cases^3,31^. The observed variability in HRD positivity across studies may be attributed to differences in assay methodologies and the thresholds used to define genomic instability. The proportions of HRD_GS-positive cases identified by AmoyDx, SOPHiA, and MG_HRD assays were 47.9%, 36.6%, and 46%, respectively, in our total study cohort. These results are slightly higher than those reported by Pepe et al. where HRD score positivity ranged between 30% to 45% for both AmoyDx and SOPHiA platforms^11^. Additionally, Romey et al. and Denkert et al. reported GIS positivity rates of 40.5% and 38.1% using the Myriad and standardized Myriad assays, respectively^30,32^. Regardless of the HRD_GS status, we observed consistent frequencies of pathogenic *BRCA1/2* mutations across all three assays ranging from 23.2 % -23.8% with an overall frequency of 23.6%. This aligns closely with the 22% prevalence reported by Landen et al study^33^.

Our study further revealed that tumors with *BRCA1/2* mutations had significantly elevated HRD genomic scores compared to those with *BRCA1/2*_wt (p < 0.0001), consistent with previous reports^32,34,35^. The median HRD scores of *BRCA1/2* mutated versus wild-type tumors were 90.3 vs. 22.2 for AmoyDx, 3.0 vs. −4.1 for SOPHiA, and 57.0 vs. 28.0 for the MG_HRD assay, respectively. Similarly, Morgan et al. and Romey et al. reported higher median GIS for Myriad assay in *BRCA1/2* mutated versus wild-type tumors (62 vs. 28; p < 0.0001)^32,34^. The observed variation in absolute median HRD scores across the assays indicates that the HRD scores are assay-specific and should not be directly compared between the platforms. Although these assays differ in scoring algorithms, genomic metrics, and assay-specific thresholds, they converge on the same biological concept that *BRCA* mutated tumors consistently harbour a higher genomic scar burden than the wild-type tumors.

A substantial proportion of tumors in our cohort were *BRCA1/2*_wt but HRD_GS-positive (28.8% overall), with frequencies ranging from 20.9% with SOPHiA, 31.6% with AmoyDx, and 29.0% with MG_HRD. This distribution is broadly consistent with previous studies, including 30% reported by Yuan et al^36^ using AmoyDx, 23% by Rojo et al^37^ using SOPHiA, 26.2% by Romey et al^32^ using Myriad, and 30.2% by Denkert et al^30^ using a standardized Myriad HRD assay. Collectively, these findings demonstrate that the frequency of this molecular subgroup of our MG_HRD assay is on par with the Myriad assay^32^. This observation highlights that a considerable subset of patients with HRD_GS positivity would be missed by *BRCA* testing alone, yet they could potentially benefit from treatment with PARPi.

Comparative analysis of *BRCA1*/*2* mutation spectra revealed a consistent predominance of truncating variants in both genes, indicating that loss-of-function is the primary mechanism of gene inactivation. Several recurrent variants were identified in the *BRCA1/2* genes; among these, the *BRCA1* c.68_69del (p.Glu23ValfsTer17) variant was the most prevalent pathogenic variant, with a frequency of 14% [97/693] detected in ovarian cancer. This hotspot mutation is also known as “185delAG” and commonly known as the “Ashkenazi Jewish population founder mutation”. This variant has also been widely reported across diverse Asian populations^38^, including the Indian population in breast and ovarian carcinoma cases. Singh et al. reported this germline variant in 3.4% (34/1,010) of patients with breast and ovarian cancer, while John et al. reported it in 24.5% of all pathogenic *BRCA1* variants [29/181] in a cohort of 921 breast and ovarian cancer cases through germline testing^39,40^. Likewise, Parween et al. observed this variant in 7.2% (8/110) of pathogenic *BRCA1* variants among patients with gynaecologic cancers, encompassing both somatic and germline tests^41^. To the best of our knowledge, this is the first study to report the *BRCA1* c.68_69del mutation exclusively in Indian patients with ovarian carcinoma using somatic *BRCA* testing methods. Collectively, these findings reinforce the prevalence of the *BRCA1* c. 68_69del variant in the Indian population and highlight its clinical significance as an important target for routine *BRCA* testing and genetic risk assessment.

Tumor cellularity is an important pre-analytical factor that can influence HRD assessment. Although a TC of approximately 30% is generally preferable for a reliable analysis, HRD testing has also been successfully performed in samples with lower TC. Romey et al. reported successful HRD analysis in 61.8% (264/427) of samples with TC<30%^32^. Furthermore, several clinically validated assays, including FoundationOne® CDx^42^, Caris HRD, SeqOne HRD solution, and CancerPrecision® (CeGaT), permit testing in samples with TC as low as 20%^10^. In our study, 512 samples with TC <30% remained evaluable for HRD classification, although HRD positivity was lower in this group than in samples with higher TC across all three assays. This finding suggests that TC may influence HRD detection while supporting the feasibility of HRD assessment in specimens with TC ≥20%. These observations are particularly relevant in real-world settings, where limited tumor material and variable tumor cellularity may otherwise restrict access to HRD testing.

HGSC was the predominant subtype in our cohort, with a significantly higher frequency than non-HGSC, consistent with prior reports. Overall, 82.5% of cases were HGSC, of which 60.2% were HRD-positive, aligning with studies demonstrating the strong association of HRD with HGSC^43^. Although in our study HRD was enriched in HGSC, it was also observed across non-HGSC subtypes. In low-grade serous carcinoma, we detected lower HRD positivity [3.8%] compared to 15% reported by Cunningham et al^44^. In clear cell carcinoma, HRD prevalence varied widely across studies, with 28% reported by Cunningham et al., 2.3% by Liu et al., while our study showed an intermediate rate of 13.7% (16/117)^44,45^. Carcinosarcoma exhibited a higher HRD frequency in our cohort (52.9%, 9/17) compared with the 25% reported by Denkert et al. This difference may reflect methodological variations between the studies, as our HRD classification incorporated both pathogenic *BRCA* mutations and genomic scar positivity, whereas Denkert et al. defined HRD positivity based on only the genomic instability score^30^. Similarly, the endometrioid carcinoma showed a higher HRD prevalence in our study (26.7%, 35/131) compared to 19% [5/26] reported by Pierson et al^46^. In contrast, mucinous carcinoma demonstrated a very low HRD positivity (2.2%), but no supporting evidence was found. Therefore, the presence of HRD in these less common histological subtypes underscores the importance of HRD profiling across these ovarian cancer subtypes, especially when they exhaust their standard therapeutic options.

*CCNE1* amplification represents an alternative oncogenic pathway associated with homologous recombination proficiency and reduced sensitivity to PARP inhibitors^7,45^. The prevalence of *CCNE1* amplification in our cohort was 7.0% (181/2596) using the MG_HRD assay, which is comparable to the 6.4% (290/4509) reported by Rojo et al.^37^ using the SOPHiA assay, but substantially lower than the 29.0% (27/93) reported by Conca et al.^35^ using the Oncomine Comprehensive Assay (OCA) Plus. These differences may reflect variations in cohort composition, assay methodology and CNV-calling criteria. These studies used different analytical definitions of amplification. For example, Conca et al. reported *CCNE1* amplification using the Median of the Absolute values of all Pairwise Differences (MAPD > 0.5) as well as the CNV p-value, whereas Rojo et al. defined amplification as more than six copies, which is relative to our assay defined amplification positive with equal or more than seven copies. In our cohort, *CCNE1* amplification was significantly enriched in *BRCA1/2*_wt tumors compared to *BRCA1/2* mutated tumors, irrespective of HRD score status. This finding is consistent with those of Rojo et al. and Conca et al., who similarly demonstrated a lower frequency of *CCNE1* amplification in *BRCA1/2* mutated tumors. Collectively, these findings support an inverse relationship between *CCNE1* amplification and pathogenic *BRCA1/2* mutations^35,37^.

Limitations of our study include lack of germline testing with paired blood samples, functional assays, and comprehensive evaluation of other non-*BRCA* HRR genes. The external orthogonal comparison cohort was limited to five samples because of logistical and cost considerations associated with centralized reference laboratory testing. Future studies should focus on prospective multicenter cohorts to validate the clinical utility of HRD testing. Further studies are needed to analyze treatment responses with longitudinal follow-up. The observation of HRD positivity in cases other than HGSC, such as carcinosarcoma and CCC, underscores the importance of non-HGOC cases for future investigations. Further exploration of resistance mechanisms, including *BRCA* reversion mutations and *CCNE1* amplification, may facilitate the development of more effective and personalized treatment strategies for patients with OC.

## Conclusions

This is the first Indian study to develop, validate, and implement an in-house HRD assay in a diagnostic laboratory. The present data evaluated across three different HRD assays enabled the assessment of HRD and *BRCA* prevalence in a large real-world ovarian cancer cohort. Our findings indicate that the MG_HRD assay is technically feasible, diagnostically reliable, and reproducible. The high level of agreement observed across platforms, including strong concordance, underscores the robustness of the MG_HRD assay as a cost-effective, flexible alternative to commercial solutions, thereby enhancing accessibility in Indian clinical settings. The findings of this study offer valuable support to clinicians and diagnostic laboratories, highlighting the use of the MG_HRD assay for assessing HRD, which can significantly benefit patients with ovarian cancer undergoing PARPi therapy, especially in lower- and middle-income countries.

## Data availability

The deidentified participant data supporting the findings of this study are available from the corresponding author upon reasonable request. Individual patient data are not publicly available due to privacy and ethical restrictions.

## Funding statement

Supported by MedGenome Labs Ltd., Bangalore, India.

## Disclosure Statement

The authors declare no conflicts of interest.

## Previous presentation

The partial data of this study entitled as “Assessing HRD and *BRCA* Status in Ovarian cancer: A real-world analysis of AmoyDx and SOPHiA assays” was presented at 4^th^ International conference on Oncopathology & Molecular Oncology organized by International Association for Integrated Oncology (IAIO) on 18^th^-19^th^ July 2026. This presentation achieved the best E-oral award.

## Acknowledgments

We thank Avantika Srishti for data curation and Merina Oliver for supporting assay validation process. We also thank the Solid Tumor Reporting Team at MedGenome Labs especially the leads Tinchan Thangamma and Himadri Baul; as well as NGS oncology lab team. We sincerely thank Dr. Rajkumar Thangarajan for his valuable input during manuscript review.

## Author Contributions

S.M.R., J.S.G., C.M.M., and V.L.R. designed the study; S.M.R., and M.P. performed literature research and conceptualized the study; S.M.R., M.P., J.S.G., C.M.M., R.G., and S.M. contributed to the study methodology, J.S.G., C.M.M., R.G., S.M., and V.L.R. planned and executed the validation; M.P., and S.M.R. analyzed and visualized the data; S.M.R., and V.L.R. supervised the study; S.M.R. and M.P. wrote the original draft; S.M.R., M.P., J.S.G., R.G., C.M.M., S.M., V.L.R., and A.K. reviewed and edited the manuscript; All other authors reviewed and approved the manuscript.

## Notes

### Competing Interest Statement

The authors have declared no competing interest.

### Author Declarations

This single-center retrospective study was approved by the Ethics committee / Institutional Review Board of the MedGenome Labs, Bangalore, India (IEC No. EC/NEW/INST/2022/3194; Proposal No. 02/2023).

